# Global Reports of Myocarditis Following COVID-19 Vaccination: A Systematic Review and Meta-Analysis

**DOI:** 10.1101/2022.03.27.22273007

**Authors:** Sirwan Khalid Ahmed, Mona Gamal Mohamed, Rawand Abdulrahman Essa, Eman Abdelaziz Ahmed Rashad, Peshraw Khdir Ibrahim, Awat Alla Khdir, Zhiar Hussen Wsu

**Affiliations:** Department of Emergency, Rania Teaching Hospital, Rania, Sulaimani, Kurdistan-region, Iraq; Department of Cardiothoracic and Vascular Surgery, Rania Medical City Hospital, Rania, Sulaimani, Kurdistan-Region, Iraq; Department of Emergency, Rania Pediatric & Maternity Teaching Hospital, Rania, Sulaimani, Kurdistan-region, Iraq; Department of Biotechnology, Institute of Science and Modern Technology, Rojava University, Qamishlo, Syria; Department of Adult Nursing, RAK Medical and Health Sciences University, Ras Al Khaimah, UAE; Department of Nursing, University of Raparin, Sulaimani, Rania, Kurdistan-region, Iraq

**Keywords:** COVID-19, Myocarditis, COVID-19 Vaccines, mRNA vaccine, pericarditis

## Abstract

In December 2020, the FDA granted emergency approval to Pfizer-BioNTech (BNT162b2) and Moderna (mRNA-1273) COVID-19 vaccines. There have been recent media reports of myocarditis after receiving COVID-19 vaccines, particularly the messenger RNA (mRNA) vaccines, causing public concern. This review summarizes information from published case series and case reports, with a strong emphasis on reporting patient and disease characteristics, investigation, and clinical outcome, to provide a comprehensive picture of the condition. Forty studies, including 147 cases, participated in this systematic review. The median age was 28.9 years; 93.9% were male and 6.1% were female. 72.1% of patients received the Pfizer-BioNTech (BNT162b2) vaccine, 24.5% of patients received the Moderna COVID-19 Vaccine (mRNA-1273), and the rest of the 3.3% received other types of vaccines. Furthermore, most myocarditis cases (87.1%) occurred after the second vaccine dose, after a median time interval of 3.3 days. The most frequently reported symptoms were chest pain, myalgia/body aches and fever. Troponin levels were consistently elevated in 98.6%. The admission ECG was abnormal in 88.5% of cases, and the left LVEF was lower than 50% in 26.5% of cases. The vast majority of patients (93.2%) resolved symptoms and recovered, and only 3 patients died. These findings may help public health policy to consider myocarditis in the context of the benefits of COVID-19 vaccination as well as to assess the cardiac condition before the choice of vaccine, which is offered to male adults. In addition, it must be carefully weighed against the very substantial benefit of vaccination.

## Introduction

International efforts to drive vaccinations are critical to restoring health and attempting economic and social recovery as the SARS-CoV-2 coronavirus (COVID-19)-caused pandemic continues [1]. The COVID-19 vaccines developed by Pfizer-BioNTech (BNT162b2) and Moderna (mRNA-1273) were granted emergency approval by the Food and Drug Administration (FDA) of the United States in December 2020. Reports of myocarditis after the COVID-19 vaccination, notably after the messenger RNA (mRNA) vaccines, have received widespread media attention in recent months, causing widespread concern among the general public [1]. Myocarditis is diagnosed in about ten to twenty people per 100,000 in the general population each year, and it has been shown to be more common in men and younger age groups [2]. Myocarditis following mRNA vaccination was first reported in Israel in April 2021, and then several case reports and case series were reported around the world.

Specifically, this report examines the current literature on myocarditis following COVID-19 vaccination, summarizing available information from previously published case reports and case series, with a strong attention on reporting patient and disease characteristics, as well as investigation and clinical outcome, in order to provide a comprehensive picture of the condition.

## Methods

### Review objectives

The main objective is to clarify the potential occurrence of myocarditis associated with COVID-19 vaccination and elaborate on the demographic and clinical characteristics of COVID-19 vaccinated individuals who develop myocarditis and how many cases have been reported in the literature.

### Protocol and Registration

The review is written in accordance with the Preferred Reporting Items for Systematic Reviews and Meta-Analyses (PRISMA) 2020 guidelines for the systematic review of available literature [3]. The protocol of the review was registered in the International Prospective Register of Systematic Reviews (PROSPERO) with ID CRD42022308997. The AMSTAR-2 checklist was also used to evaluate this study, and it was found to be of high quality [4]. This review article didn’t need to get ethics approval.

### Search strategy

A comprehensive search of major electronic databases (PubMed and Google Scholar) was conducted on February 10, 2022, to locate all publications. The AND operator was used to connect two of the most important concepts in the search terminology (“COVID-19” AND “Myocarditis”). (“Myocarditis” and “COVID-19” OR “SARS-CoV-2” OR “Coronavirus Disease 2019” OR “severe acute respiratory syndrome coronavirus 2” OR “coronavirus infection” OR “2019-nCoV” AND “vaccine, vaccination, OR vaccine” were used in the search. To make sure the search was completed, we checked the references of all relevant papers.

### Eligibility criteria

All case series and case reports on post-COVID-19 vaccine myocarditis in humans were included. Individuals who develop myocarditis after receiving the COVID-19 vaccine, regardless the type of vaccine and dose. The references of the relevant articles will also be reviewed for additional articles that meet the inclusion criteria. Narrative and systematic reviews, original and unavailable data papers were excluded in this review. Moreover, articles other than English were excluded in this review.

### Data extraction and selection process

PRISMA 2020 was used to guide every step of the data extraction process from the original source. The Rayyan website was used by two independent authors (SKA and RAE) to screen abstracts and full-text articles based on inclusion and exclusion criteria [5]. The third author (RAE) resolved any discrepancies between the two independent authors. Microsoft Excel spreadsheets were used to collect the necessary information from the extracted data. Author names, year of publication, age, gender, type of COVID-19 vaccine, dose, days to symptoms onset, symptoms, troponin level, LVEF 50% or LVEF > 50%, ECG, length of hospital stay/days, treatment, and outcomes were extracted from each study.

### Critical appraisal

To assess the quality of all included studies, we used the Joanna Briggs Institute’s critical appraisal tool for case series and case reports [6]. Each article was evaluated by two different authors (SKA and RAE), each of whom worked independently. Paper evaluation disputes were resolved either through discussion or by a third author (RAE) depending on the circumstances. Articles with an average score of 50 percent or higher were included in the data extraction process. The AMSTAR 2 criteria [4]. were used to evaluate the results of our systematic review. The AMSTAR 2 tool assigned a “medium” rating to the overall quality of our systematic review.

### Data synthesis and analysis

All the articles included in the current systematic review were analysed, and the data was extracted and pooled. This included (authors’ names, year of publication; gender; type of COVID-19 vaccine, dose, days to symptoms onset, troponin level, LVEF below or above 50%, ECG, length of hospital stay/days; treatment and outcomes). We gathered this data from the results of eligibility studies. COVID-19 vaccine recipients who developed myocarditis were included in the study.

## Results

### Selection of studies

When we searched the major databases (PubMed and Google Scholar) on February 10, 2022, we discovered 2712 articles that were relevant to our search criteria. A citation manager tool (Mendeley) was then used to organize the references, and 283 articles were automatically removed because they contained duplicate content. Next, the titles, abstracts, and full texts of 2429 articles were checked for accuracy, and 2365 articles were rejected because they did not meet the criteria for inclusion. Besides that, 64 articles were submitted for retrieval, but twenty-four were rejected because they did not meet our inclusion requirements. The current systematic review was limited to 40 articles in total **(Fig 1)**. The details of case reports and case series are shown in **(Table 1)**.

**Table 1:** Characteristics and outcomes of patients with myocarditis related to COVID-19 vaccine.

| Author/Y<br>ear of<br>publicati<br>on | Cou<br>ntry | Age | Gen<br>der | Type of<br>COVID-19<br>vaccine | Dose | Days to<br>symptom<br>onset | Symptoms | Troponi<br>n level | LVEF<br><50% or<br>LVEF<br>>50% | Electroc<br>ardiogr<br>am<br>(ECG) | Treatment | Length<br>of<br>hospital<br>stay<br>(days) | Outco<br>me |
| --- | --- | --- | --- | --- | --- | --- | --- | --- | --- | --- | --- | --- | --- |
| Abu<br>Mouch et<br>al 2021<br>[24] | Israel | 6 cases<br>mean<br>age 22<br>years | All<br>of<br>them<br>were<br>male | BNT162b<br>2 | 2<sup>nd</sup> in 5<br>cases<br>and 1<sup>st</sup><br>in one<br>case | Mean 4.5<br>days | Chest<br>pain/discomfort | Elevated<br>in all<br>cases | LVEF<br>>50% in all<br>cases | Abnorm<br>al in all<br>cases | NSAIDs and<br>colchicine | Mean<br>5.6<br>days | Recover<br>ed |
| Marshall<br>et al 2021<br>[25] | USA | 7 cases<br>mean<br>age<br>16.7<br>years | All<br>of<br>them<br>were<br>male | BNT162b<br>2 | 2<sup>nd</sup> | Mean 2.57<br>days | Chest pain | Elevated<br>in all<br>cases | LVEF<br>>50% in 6<br>cases<br>and<br>LVEF<br><50% in<br>one case | Abnorm<br>al in all<br>cases | NSAIDs,<br>IVIg, IV<br>methylprednis<br>olone, PO<br>prednisone,<br>famotidine,<br>aspirin | Mean<br>11.57<br>days | Recover<br>ed |
| D'Angelo<br>et al 2021<br>[26] | Italy | 30<br>years | Male | BNT162b<br>2 | 1<sup>st</sup> | 21 days | dyspnea,<br>constrictive<br>retrosternal pain,<br>nausea,<br>and profuse<br>sweating | Elevated | LVEF<br>>50% | Abnorm<br>al | Bisoprolol,<br>aspirin, and<br>prednisolone | 7 days | Recover<br>ed |
| Nassar et<br>al 2021<br>[27] | USA | 70<br>years | Fema<br>le | Ad26.CO<br>V2. S | 1<sup>st</sup> | 2 days | The patient<br>arrived at the<br>emergency<br>department in<br>severe<br>respiratory<br>distress | Elevated | LVEF<br>>50% | Abnorm<br>al | vasopressors<br>and antibiotic<br>therapy | 8 days | Died |
| Kim et al<br>2021 [28] | USA | 4 cases<br>mean<br>age<br>38.25<br>years | 3<br>male<br>s and<br>1<br>fema<br>le | mRNA-1273 in 2<br>cases<br>And<br>BNT162b<br>2 in 2<br>cases | 2<sup>nd</sup> | Mean 2.75<br>days | Chest pain | Elevated<br>in all<br>cases | LVEF<br>>50% in 3<br>cases and<br>LVEF<br><50% in<br>one case | Abnorm<br>al in all<br>cases | Corticosteroid<br>s NSAIDs and<br>colchicine | Mean<br>2.5<br>days | Recover<br>ed |
| Montgom<br>ery et al<br>2021 [10] | USA | 23<br>cases<br>mean<br>age 25<br>years | All<br>of<br>there<br>were<br>male | BNT162b<br>2 in 7<br>cases and<br>16 cases<br>mRNA-1273 | 2<sup>nd</sup> in<br>20<br>cases<br>and<br>1<sup>st</sup> in 3<br>cases | Mean 2<br>days | Chest pain | Elevated<br>in all<br>cases | LVEF<br><50% in 4<br>cases and<br>LVEF<br>≥50% in 19<br>cases | Abnorm<br>al in 19<br>cases<br>and<br>normal<br>in 4<br>cases | All patients<br>received brief<br>supportive<br>care | Mean<br>7 days | 16 cases<br>were<br>fully<br>recover<br>ed and 7<br>cases<br>under<br>follow-up |
| Verma et<br>al 2021<br>[29] | USA | 2 cases<br>(45,<br>42)<br>years<br>Mean<br>age<br>43.5<br>years | 1<br>male<br>and<br>1<br>fema<br>le | BNT162b<br>2- in 1<br>case and 1<br>case<br>mRNA-1273 | 1<sup>st</sup> in<br>one<br>case<br>and 2<sup>nd</sup><br>in<br>anothe<br>r case | Mean 12<br>days | Chest pain,<br>dyspnea and<br>dizziness, | Elevated<br>in all<br>cases | LVEF<br>>50% in all<br>cases | Abnorm<br>al in all<br>cases | intravenous<br>diuretics,<br>methylprednis<br>olone,<br>lisinopril,<br>spironolacton<br>e, and<br>metoprolol<br>succinate). | 7 days | female<br>case<br>recover<br>ed and<br>male<br>case<br>died |
| Rosner et<br>al 2021<br>[30] | USA | 7 cases<br>Mean<br>age<br>27.42<br>years | All<br>of<br>them<br>were<br>male | BNT162b<br>2 in 5<br>cases, one<br>case<br>mRNA-1273 and<br>one case<br>Ad26.CO<br>V2. S | 2<sup>nd</sup> in 6<br>cases<br>and 1<sup>st</sup><br>in one<br>case | Mean 3.85<br>days | Chest pain | Elevated<br>in all<br>cases | LVEF<br>>50% in 6<br>cases and<br>LVEF<br><50% in<br>one case | Abnorm<br>al in 6<br>cases<br>and<br>normal<br>in one<br>case | β-blocker and<br>anti-inflammatory<br>medication | Mean<br>2.85<br>days | Recover<br>ed |
| Dionne et al 2021 [31] | USA | 15 cases mean age 15 years | 14 cases male and one case female | BNT162b 2 in all cases | 2<sup>nd</sup> in all cases | Mean 3 days | Chest pain, fever, myalgia, headache | Elevated in all cases | Mean LVEF <50% in all cases | Abnormal in all cases | β-blocker therapy. | Mean 2 days | Recovered |
| García et al 2021 [32] | Mexico | 39 years | Male | BNT162b 2 | 2<sup>nd</sup> | ¼ day | Chest pain | Elevated | LVEF >50% | Abnormal | anti-inflammatory medication | 6 days | Recovered |
| Dickey et al 2021 [33] | USA | 6 cases mean age 27 years | All of them were male | BNT162b 2 in 5 cases and one case mRNA-1273 | 2<sup>nd</sup> | Mean 3.33 days | chest pain, chills, myalgia, malaise, headache and fever | Elevated in all cases | LVEF >50% in 3 cases and LVEF <50% in 3 cases | Abnormal in 5 cases and normal in one case | Unknown | Unknown | Recovered |
| Tano et al 2021 [34] | USA | 8 cases mean age 16.61 years | All of them were male | BNT162b 2 in all cases | 2<sup>nd</sup> in 7 cases and 1<sup>st</sup> in one case | Mean 2.37 days | Chest pain, fatigue, abdominal pain, fever, shortness of breath | Elevated in all cases | LVEF >50% in all cases | Abnormal in 6 cases and normal in 2 cases | NSAIDs | Mean 2.36 days | Recovered |
| Larson et al 2021 [35] | USA and Italy | 8 cases mean age 31.62 years | All of them were male | BNT162b 2 in 5 cases and 3 cases mRNA-1273 | 2<sup>nd</sup> in 7 cases and 1<sup>st</sup> in one case | Mean 2.75 days | Chest pain, myalgia, fever, chills, shortness of breath and cough | Elevated in all cases | LVEF >50% in 6 cases and LVEF <50% in 2 cases | Abnormal in 7 cases and normal in 1 case | NSAIDs, colchicine and prednisone | Unknown | Recovered |
| Deb et al 2021 [36] | USA | 67 years | Male | mRNA-1273 | 2<sup>nd</sup> | ¼ day | Nausea, orthopnea, fatigue | Elevated | LVEF >50% | Normal | intravenous furosemide, bronchodilators | **2 days** | **Recovered** |
| Abbate et al 2021 [37] | USA | 2 cases mean age 30.5 years | One male and one female | BNT162b 2 in all cases | 2<sup>nd</sup> in one case and 1<sup>st</sup> in second case | Mean 5.5 days | Fever, cough, chest pain, nausea and vomiting | Unknown | LVEF <50% in all cases | Abnormal in all cases | Prednisone | 73 days for one case | One case died and one recovered |
| Muthukumar et al 2021 [38] | USA | 52 years | Male | mRNA-1273 | 2<sup>nd</sup> | 1 day | Chest pain, fevers, shaking chills, myalgias, and headache | Elevated | LVEF <50% | Abnormal | low-dose lisinopril and carvedilol | 4 days | Recovered |
| Isaak et al 2021 [39] | Germany | 15 years | Male | BNT162b 2 | 2<sup>nd</sup> | 1 day | fever, myalgia | Elevated | LVEF <50% | Abnormal | Unknown | 7 days | Recovered |
| Cereda et al 2021 [40] | Italy | 12 years | Male | BNT162b 2 | 2<sup>nd</sup> | 1 day | Chest pain | Elevated | LVEF >50% | Abnormal | Bisoprolol and ramipril | 7 days | Recovered |
| Watkins et al 2021 [41] | USA | 20 years | Male | BNT162b 2 | 2<sup>nd</sup> | 2 days | Chest pain and shortness of breath | Elevated | LVEF >50% | Abnormal | Colchicine, metoprolol, ibuprofen | Unknown | Recovered |
| Chamling et al 2021 [42] | Germany | 3 cases mean age 37.66 years | 2 males and 1 female | BNT162b 2 in 2 cases, and ChAdOx1 nCoV-19 in 1 case | 1<sup>st</sup> in 2 cases and 2<sup>nd</sup> in 1 case | Mean 7 days | Chest pain | Elevated in 2 cases and not elevated in 1 case | LVEF >50% in all cases | Abnormal in 2 cases and normal on 1 case | Unknown | Unknown | Recovered |
| Mansour et al 2021 [43] | USA | 2 cases mean age 23 years | 1 male and 1 female | mRNA-1273 in all cases | 2<sup>nd</sup> in all cases | 1 day | Chest pain, fever, | Elevated in all cases | LVEF >50% in all cases | Abnormal in all cases | metoprolol | Mean 2 days | Recovered |
| Levin et al 2021 [44] | Israel | 7 cases mean age 20.42 years | All of them were male | BNT162b 2 in all cases | 2<sup>nd</sup> in all cases | Mean 7 days | Chest pain, fatigue, fever and headache | Elevated in all cases | LVEF >50% in 5 cases LVEF <50% in 2 cases | Abnormal in all cases | Colchicine, Ibuprofen, Bisoprolol, and Ramipril | Mean 2.5 days | Recovered |
| Schauer et al 2021 [45] | USA | 13 cases mean age 15.07 years | 12 male and 1 female | BNT162b 2 in all cases | 2<sup>nd</sup> in all cases | Mean 2.76 days | chest pain, shortness of breath, fever and myalgia | Elevated in all cases | LVEF >50% in 11 cases LVEF <50% in 2 cases | Abnormal in 9 cases and normal in 4 cases | NSAIDs | Mean 2 days | Recovered |
| Shumkova et al 2021 [46] | Poland | 23 years | Male | BNT162b 2 | 2<sup>nd</sup> | 1 day | chest pain, shortness of breath and fever | Elevated | LVEF >50% | Abnormal | Aspirin and methylprednisolone | 6 days | Recovered |
| Minocha et al 2021 [47] | USA | 17 years | Male | BNT162b 2 | 2<sup>nd</sup> | 2 days | chest pain | Elevated | LVEF >50% | Abnormal | NSAIDs | 6 days | Recovered |
| Hasnie et al 2021 [48] | USA | 22 years | Male | mRNA-1273 | 1<sup>st</sup> | 3 days | Chest pain | Elevated | LVEF >50% | Abnormal | Aspirin and colchicine | 2 days | Recovered |
| Starekova et al 2021 [49] | USA | 5 cases mean age 25.2 years | 4 males and 1 female | BNT162b 2 in 3 cases and mRNA-1273 in 2 cases | 2<sup>nd</sup> in all cases | Mean 2.6 days | Chest pain, fatigue, nausea, fever, chills and myalgia | Elevated in all cases | LVEF >50% in 4 cases LVEF <50% in 1 case | Abnormal in 4 cases and normal in 1 case | Unknown | Unknown | Recovered |
| Koizumi et al 2021 [50] | Japan | 2 cases mean age 24.5 years | All of them were male | mRNA-1273 in all cases | 2<sup>nd</sup> in all cases | Mean 2.5 days | Chest pain | Elevated in all cases | LVEF >50% in all cases | Abnormal in all cases | NSAIDs | Mean 4 days | Recovered |
| McLean et al 2021 [51] | USA | 16 years | Male | BNT162b 2 | 2<sup>nd</sup> | 1 day | Chest pain | Elevated | LVEF >50% | Abnormal | NSAIDs | 7 days | Recovered |
| Riedel et al 2021 [52] | Brazil | 47 years | Male | Sinovac COVID-19 vaccine in activated (China) | 2<sup>nd</sup> | Unknown | Chest pain Cough and myalgia | Elevated | LVEF <50% | Abnormal | Unknown | Unknown | Recovered |
| In-Cheol et al 2021 [53] | Korea | 24 years | Male | BNT162b 2 | 2<sup>nd</sup> | 1 day | Chest pain | Elevated | LVEF >50% | Abnormal | Unknown | 5 days | Recovered |
| Nguyen et al 2021 [54] | Germany | 20 years | Male | mRNA-1273 | 1<sup>st</sup> | 1 day | Chest pain, fatigue and myalgia | Elevated | LVEF >50% | Abnormal | Unknown | Unknown | Recovered |
| Azdaki et al 2021 [55] | Iran | 70 years | Male | ChAdOx1 nCoV-19. | 1<sup>st</sup> | 3 days | Syncope | Elevated | LVEF >50% | Abnormal | magnesium sulfate | 7 days | Recovered |
| Sokolska et al 2021 [56] | Poland | 21 years | Male | BNT162b 2 | 1<sup>st</sup> | 3 days | Chest pain | Elevated | LVEF >50% | Abnormal | Unknown | Unknown | Recovered |
| Patel et al 2021 [57] | USA | 5 cases mean age 23.2 years | All of them were male | BNT162b 2 in 4 cases and mRNA-1273 in 1 case | 2<sup>nd</sup> in 4 cases and 1<sup>st</sup> in 1 case | Mean 2.2 days | Chest pain, dyspnea, nausea, headache and chills | Elevated in all cases | LVEF >50% in all cases | Abnormal in all cases | Colchicine, Ibuprofen and aspirin | Mean 1.8 days | Recovered |
| Kim et al 2021 [58] | Korea | 29 years | Male | BNT162b 2 | 2<sup>nd</sup> | 1 day | Chest pain | Elevated | LVEF >50% | Normal | corticosteroids and NSAIDs. | 7 days | Recovered |
| Ehrlich et al 2021 [59] | Germany | 40 years | Male | BNT162b 2 | 1<sup>st</sup> | 6 days | chest pain and shortness of breath, and fever | Elevated | LVEF <50% | Abnormal | Aspirin, heparin, beta-blocker and a mineralocorticoid antagonist | 2 days | Recovered |
| Schmitt et al 2021 [60] | France | 19 years | Male | BNT162b 2 | 2<sup>nd</sup> | 3 days | Chest pain and dyspnea | Elevated | LVEF >50% | Abnormal | Unknown | 1 day | Recovered |
| Kadwalwala et al 2021 [61] | USA | 38 years | Male | mRNA-1273 | 1<sup>st</sup> | 2 days | Chest pain, fatigue and fever | Elevated | LVEF <50% | Abnormal | Methylprednisolone, lisinopril, and spironolactone | 6 days | Recovered |
| Azir et al 2021 [62] | USA | 17 years | Male | BNT162b 2 | 2<sup>nd</sup> | 1 day | Chest pain | Elevated | LVEF >50% | Abnormal | aspirin and sublingual nitroglycerin | 1 day | Recovered |

**Fig 1:**
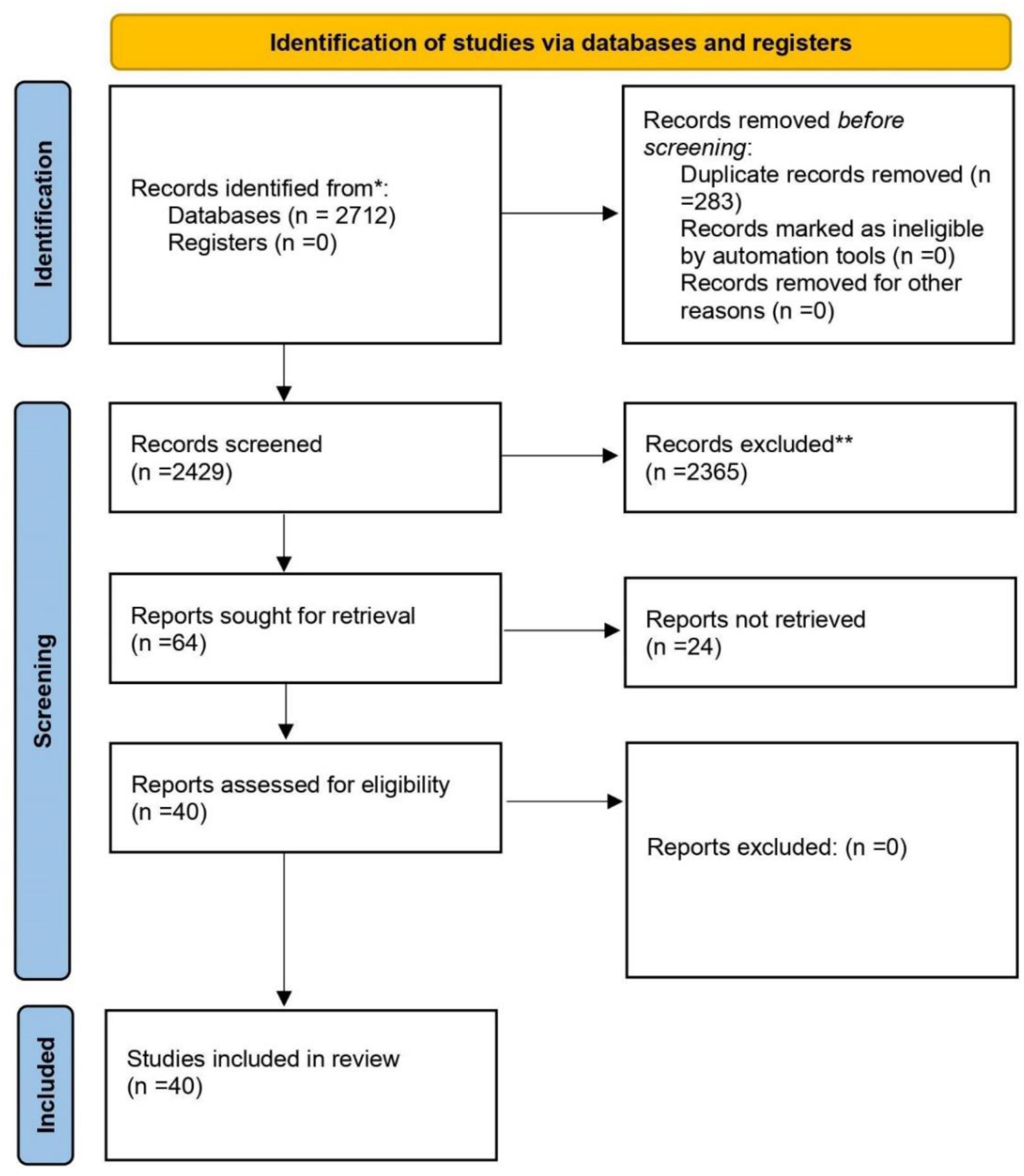
PRISMA flow-diagram

### Characteristics of the included studies

Overall, forty studies, including 147 cases each, from the United States, Italy, Israel, Germany, Poland, France, Korea, Brazil, Japan, Mexico, and Iran participated in this systematic review. The median age was 28.9 years; 93.9% were male and 6.1% were female. 72.1% of patients received the Pfizer-BioNTech (BNT162b2) vaccine, 24.5% of patients received the Moderna COVID-19 Vaccine (mRNA-1273), and the rest of the 3.3% received other types of vaccines (Johnson & Johnson, AstraZeneca, Sinovac).

The vast majority of cases are from the United States. All patients were diagnosed with myocarditis or myopericarditis following COVID-19 vaccination, regardless of the type of vaccine and dose.

Furthermore, most myocarditis cases (87.1%, n = 128) occurred after the second vaccine dose, after a median time interval of 3.3 days. The most frequently reported symptoms were chest pain (100% n = 147), fever (46.9% n = 69), myalgia/body aches (54.4% n = 80), and also variable reports of viral prodromes such as chills, headaches, and malaise. Troponin levels were consistently elevated in 98.6% (n = 145) of the cases where they were reported, consistent with myocardial injury. The admission electrocardiogram (ECG) was abnormal in 88.5% (n = 130) of cases, and the left ventricular ejection fraction (LVEF) was lower than 50% in 26.5% (n = 39) of cases. The median length of hospital stay was 5.2 days in 127 patients but unknown in 20 patients. The vast majority of patients (93.2%) (n = 137) resolved symptoms and recovered, and only 3 patients died (**Table 2**).

**Table 2:** Summary of pooled data from included research papers have been reported in the literature (n = 147)

|  |  |
| --- | --- |
| **Age** | Mean age - 28.94 years |
| **Gender (n) %** | Male – 138 (93.9%) |
|  | Female – 9 (6.1%) |
| **Type of COVID-19 vaccine (n) %** | Pfizer-BioNTech (BNT162b2) – 106 (72.1%) |
|  | Moderna COVID-19 Vaccine (mRNA-1273) - 36 (24.5%) |
|  | Janssen (Johnson & Johnson) ( Ad26.COV2. S) – 2 (1.3%) |
|  | Oxford, AstraZeneca COVID-19 vaccine ChAdOx1 nCoV-19 – 2 (1.3%) |
|  | Sinovac COVID-19 vaccine in activated – 1 (0.7%) |
| **Dose (n) %** | First dose – 19 (12.9%) |
|  | Second dose – 128 (87.1%) |
| **Days to symptom onset** | Median 3.32 days |
| **Symptoms (n) %** | Chest pain - 147 (100%) |
|  | Fever – 69 (46.9%) |
|  | Myalgia/body aches – 80 (54.4) |
|  | Shortness of breath – 11 (7.5%) |
| **Troponin level (n) %** | Elevated – 145 (98.6%) |
|  | Not elevated – 1 (0.6%) |
|  | Unknown – 1 (0.6%) |
| **LVEF (n) %** | LVEF >50% - 108 (73.5%) |
|  | LVEF <50% - 39 (26.5%) |
| **Electrocardiogram (ECG) (n) %** | Abnormal – 130 (88.5%) |
|  | Normal – 17 (11.5%) |
| **Length of hospital stay (days)** | Median 5.2 days in 127 patients |
|  | Unknown in 20 patients |
| **Outcome (n) %** | Recovered – 137 (93.2%) |
|  | Under follow-up – 7 (4.7%) |
|  | Died – 3 (2%) |

## Discussion

The current systematic review summarized evidence from the original case reports and case series that explored the development of myocarditis after COVID-19 vaccination. Throughout the selected studies, most of the participants were male, from the USA, and their mean age 28.94 was years old. The mechanism of vaccine-induced myocarditis is not known but may be related to the active pathogenic component of the vaccine and specific human proteins, which could lead to immune cross-reactivity resulting in autoimmune disease, which is one cause of myocarditis [7–10]. The occurrence of myocarditis in men may be related to sex hormone variations, as testosterone hormone suppresses anti-inflammatory immune cells while promoting more aggressive T helper cells [7,11].

These findings were matched with Oster et al (2022) [12], who found the incidence rate of myocarditis among vaccinated male people was similar to that seen in typical cases of myocarditis and there was a strong male predominance for both conditions [13]. Fatima et al (2022) [7] found most patients who developed myocarditis were males. Moreover, Patone et al (2022) [14] mentioned that the incidence of myocarditis was among England males younger than 40 years old. Similarly, a systematic review study found that the Incidence of myocarditis following mRNA vaccines is low but probably highest in males aged 12-29 years old [15].

Another important finding in the current systematic review is that most of the participants received Pfizer-BioNTech (BNT 162b2) followed by the Moderna CVID-19 vaccine (mRNA-1273), and most of the cases who complained of myocarditis received two doses of the vaccine. This indicates that mRNA vaccines are associated with a higher risk of developing myocarditis than viral vector vaccines, including Janssen, Oxford, and Sinovac. Bozkurt et al (2021) [2], have assumed that autoantibody generation could attack cardiac myocytes in response to mRNA vaccine, which increases the risk.

This finding is matched with Patone (2022) [14], who mentioned the risk of myocarditis increased within a week of receiving the first dose of both adenovirus and mRNA vaccines and after the second dose of mRNA vaccine. Oster et al (2022) [12] concluded that the risk of myocarditis after the mRNA vaccine was increased after the second dose in adolescents and young males. On the other hand, Simone et al (2021) [16] concluded that there is no relationship between COVID-19 mRNA vaccination and post vaccination myocarditis.

The findings extend these observations, which include the median onset of symptoms after vaccine administration was 3.32 days. The most common symptoms are chest pain, followed by myalgia/ body aches and fever. These findings matched with Pillay et al (2021) [15], who reported in a systematic review observation that the majority of myocarditis cases had a short symptoms onset of 2 to 4 days after a second dose, and the majority presented with chest pain. These findings matched with Oster et al (2022) [12], who mentioned myocarditis was diagnosed within days of vaccination.

In patients with severe myocarditis, the diagnosis is often established by heart biopsy. In patients with mild myocarditis, the diagnosis is based on compatible clinical findings and confirmed by elevated levels of blood markers or an electrocardiogram (ECG) indicative of cardiac injury, with the presence of new abnormalities on echocardiography or cardiac MRI [17].

Cardiac-specific investigations revealed that troponin levels were elevated in almost all of the cases, consistent with myocardial injury, which is associated with autoimmune processes matched with vaccine protein and the case immune system.

In the same line with Lee et al. (2022) [1], a systematic review to investigate myocarditis following COVID-19 Vaccination in October 2020–October 2021, mentions that all reported cases have an elevated troponin level in keeping with myocardial injury.

In our study, less than one third of cases had left ventricle ejection friction (LVEF) was less than 50%. Compared to patients with COVID-19 illness, patients with vaccine associated myocarditis had a higher LVEF.

This finding is consistent with the findings of Fronza et al. (2022) [18], who investigated myocardial injury patterns at MRI in COVID-19 Vaccine and discovered that more than half of the cases had more than 50% LVEF. Also, Shiyovich et al. (2022) [19], who analyzed myocarditis following the third (Booster) dose of COVID-19 vaccination found that the mean left ventricular ejection fraction was 61 ± 7% (range 53–71%) and regional wall motion abnormalities were present in one of the patients only. Global T1 values were increased in one (25%) of the patients, while focal values were increased in 3 (75%) of the patients. Global T2 values were increased in one (25%) of the patients, while focal values were increased in all of the patients (100%). Global ECV was increased in 3 (75%) of the patients, while focal ECV was increased in all the patients (100%). LGE was present in all the patients.

In our systematic review and meta-analysis study, 88.5% of cases had abnormal changes in the electrocardiogram (ECG) result, regardless of the vaccine type.

Vidula et al. (2021) [20] support our findings by reporting two patients with clinically suspected myocarditis who presented with acute substernal chest pain and/or dyspnea after receiving the second dose of the vaccine and were found to have diffuse ST elevations on electrocardiogram (ECG), elevated cardiac biomarkers and inflammatory markers, and mildly reduced left ventricular (LV) function on echocardiography.

Also, Puchalski et al. [21] reported the findings of a case series regarding COVID-19-Vaccination-Induced Myocarditis in Teenagers. Electrocardiogram (ECG) patterns varied, but in all cases, characteristic features of acute myocardial injury, including ST segment elevation or depression, and repolarization time abnormalities, were present.

Management of myocarditis remains largely supportive and is based on the restoration of hemodynamic stability and the administration of guideline-directed heart failure and arrhythmia treatment. Patients with preserved ventricular function and non-severe features were often treated with colchicine or non-steroidal anti-inflammatory drugs. According to our findings, all cases were treated with NSAIDs, beta blockers, calcium channel blockers, and/or diuretics. The median length of hospital stay was 5.2 days in 127 patients, and the vast majority of patients resolved symptoms and recovered, and only 3 patients died.

This finding broadly supports the work of other studies in this area. Woo et al [22] reported that many patients who received anti-inflammatory agents such as NSAIDs, colchicine, steroids, and intravenous immunoglobulin recovered without further medical treatment, with a hospital stay lasting 3-6 days.

In accordance with the present results, previous studies have demonstrated that almost all of the cases experienced a prompt recovery with no residual cardiac dysfunction. The median length of stay for all myocarditis cases was around 2–3 days, with a range of 2–10 days [23].

## Conclusion

In conclusion, these findings may help public health policy to consider myocarditis in the context of the benefits of COVID-19 vaccination as well as to assess the cardiac condition before the choice of vaccine, which is offered to male adults. In addition, it must be carefully weighed against the very substantial benefit of vaccination. Moreover, further research is required to assess the long-term consequences and other risk factors following immunization, specifically the mRNA-1273 vaccine.

## Data Availability

All data produced in the present work are contained in the manuscript

## Author Agreement Statement

We declare that this manuscript is original, has not been published before and is not currently being considered for publication elsewhere. We confirm that the manuscript has been read and approved by all named authors and that there are no other persons who satisfied the criteria for authorship but are not listed. We confirm that the order of authors listed in the manuscript has been approved by all of us. We understand that the Corresponding Author is the sole contact for the Editorial process. He is responsible for communicating with the other authors about progress, submissions of revisions and final approval of proofs.

## Data availability Statement

All relevant data are within the manuscript and its supporting information files.

## Authors’ contributions

Conception and design SKA acquisition of data SKA, RAE, MGM, EAA analysis and interpretation of data SKA, MGM, RAE, EEA, drafting of the manuscript SKA, RAE, MGM, EAA, PKI critical revision of the manuscript for important intellectual content statistical analysis SKA, MGM, RAE, EEA, PKI, AAK, ZHW administrative SKA, MGM, EAA technical SKA, AAK, ZHW, supervision SKA, RAE and all authors approving final draft.

## Provenance and peer review

Not commissioned, externally peer-reviewed

## Acknowledgments

Not applicable

